# Associations of joint swelling, joint stiffness, and joint pain with physical activity in first-degree relatives of patients with rheumatoid arthritis

**DOI:** 10.1101/2020.06.13.20130526

**Authors:** Jan M. Hughes-Austin, Joachim H. Ix, Samuel R. Ward, Michael H. Weisman, James R. O’Dell, Ted R. Mikuls, Jane H. Buckner, Peter K. Gregersen, Richard M. Keating, M. Kristen Demoruelle, Kevin D. Deane, V. Michael Holers, Jill M. Norris

**Author notes:** ^*^**CORRESPONDING AUTHOR** Jan Hughes-Austin, PT, PhD, University of California, San Diego, Department of Orthopaedic Surgery, 9500 Gilman Drive, Mail Code 0863, La Jolla, CA 92093.

## Abstract

**Background:** Physical activity (PA) in pre-clinical rheumatoid arthritis (RA) is associated with lower RA risk and disease severity. Yet, PA in RA patients is less than in counterparts without RA, which can be attributed partly to symptoms of inflammatory arthritis. Therefore, we investigated whether joint swelling, stiffness, or pain were associated with PA in first-degree relatives (FDRs) of RA patients, a population at higher risk for future RA.

**Methods:** We evaluated associations of joint stiffness, joint swelling, and joint pain with PA time in 268 FDRs with ≥2 visits over an average 1.2 years. Clinicians confirmed joint swelling. Participants self-reported joint stiffness and/or pain. PA during a typical 24-hour day was quantified via questionnaire, weighted to reflect metabolic expenditure, where 24 hours was the minimum PA time. Linear mixed models evaluated associations between symptoms and change in PA over time, adjusting for age, sex, race, body mass index, smoking, and RA-related autoantibodies.

**Results:** Average weighted PA time was 37±7 hours. In cross-sectional analysis, PA time was 1.3±0.9 hours higher in FDRs reporting joint pain (p=0.15); and 0.8±1.6 and 0.4±1 hours lower in FDRs with joint swelling (p=0.60) and stiffness (p=0.69), respectively. Longitudinally, adjusting for baseline PA time, baseline symptoms were not significantly associated with changes in PA time. However, on average over time, joint stiffness and pain were associated with lower PA time (p_interaction_=0.0002, p_interaction_=0.002), and joint swelling was associated with higher PA time (p_interaction_<0.0001)

**Conclusion:** Baseline symptoms did not predict future PA time, but on average over time, joint symptoms influenced PA time.

## INTRODUCTION

Rheumatoid arthritis (RA) is a chronic systemic inflammatory disease that leads to pain, joint destruction, disability, and reduced life expectancy. Physical activity is encouraged in RA to facilitate joint mobility, physical function, and mitigate cardiovascular disease;^1^ yet it is significantly less than in counterparts without RA,^2, 3^ where muscle strength just exceeds thresholds needed to maintain activities of daily living.^2, 3^ Several indicators of RA disease activity, such as joint flares,^4^ pain level and fatigue, reduced mobility, and stiffness,^2, 5^ have been inversely associated and recognized as barriers to physical activity engagement.^2, 4, 5^

Physical activity prior to RA has been associated with lower risk for RA, a milder disease course, and less severe joint destruction.^6, 7^ Thus, physical activity could extend the time until RA becomes clinically manifest, and presents a key opportunity to intervene in the pre-clinical period. Further, beyond prevention, if individuals are diagnosed with RA at a time when they have more physical activity, ability, and strength, it may lead to them maintaining physical activity and independent living during the course of the disease.

Pre-clinical RA is characterized by the development of autoantibodies, circulating inflammatory markers, and signs and symptoms of inflammatory joint disease.^8-10^ It is not known in people at risk for RA who exhibit pre-clinical RA features whether inflammatory joint signs and symptoms are associated with more or less physical activity time. Therefore, we hypothesized *a priori* that inflammatory joint symptoms such as joint swelling, joint stiffness, and joint pain would be associated with less physical activity time. Using first-degree relatives (FDRs) of RA patients from the Studies of the Etiology of Rheumatoid Arthritis (SERA), a population without RA but at increased risk for RA, we sought to determine whether joint swelling, joint stiffness, and joint pain are associated with physical activity time at baseline and over time.

## METHODS

### Studies of the Etiology of Rheumatoid Arthritis (SERA)

Founded in 2003, SERA is a multi-center study designed to examine the role of environmental and genetic factors in the development and progression of RA-related autoimmunity, and to explore pre-clinical immunological changes and pathophysiological processes in the absence of treatments or secondary complications of active RA disease.^9^ The SERA cohort consists of FDRs [parent, sibling, or offspring] of probands with RA, who are selected for this prospective study because of higher RA risk. FDRs were recruited through probands [identified from academic centers, Veterans’ hospitals and rheumatology clinics or through responses to advertising] from 6 sites across the US [Denver, CO, Los Angeles, CA, Seattle, WA, Manhassett, NY, Omaha, NE, and Chicago, IL]. FDRs were eligible to participate if they did not have an RA diagnosis at the time of their initial visit, as defined by 1987 ACR Criteria,^11^ and were ≥18 years old. At research visits, FDRs completed disease and exposure assessment questionnaires, underwent a standardized interview and 68-count joint examination by a trained study clinician, and had blood drawn. FDRs positive for any RA-related autoantibody at visits were seen annually while autoantibody negative FDRs were seen every two years. Institutional Review Boards at each of the SERA sites approved this study, and all participants provided written informed consent.

### Study Population

In SERA, physical activity questions were not routinely asked until 2010, so we used the first time an FDR answered this question, between 2010 and 2014, as their “baseline” visit. There were 878 FDRs seen in clinic within this 4-year period who had complete information regarding physical activity and the presence of joint swelling, joint stiffness, or joint pain. Of these 878 FDRs, 268 FDRs had ≥ 2 clinic visits, which allowed us to also investigate changes in PA over time. There were no significant differences between this subset of 268 FDRs and the larger cohort of 878 FDRs with regards to demographics and clinical characteristics. These 268 FDRs with multiple clinic visits comprised our analysis dataset.

### RA-Related Inflammatory Joint Swelling or Symptoms

Trained clinicians, blinded to autoantibody status, confirmed the presence of joint swelling using a 68-count joint examination during clinic visits. Additional inflammatory joint symptoms involving the same joints were indicated by the presence of self-reported joint stiffness and pain during the same clinic visit. Consistent with Sparks and colleagues, we included joints at RA-specific sites according to the ACR/EULAR 2010 classification for RA, i.e., any of the metacarpophalangeal (MCP), proximal interphalangeal (PIP), or metatarsophalangeal (MTP) joints, wrist, as well as the elbow due to its relative specificity for RA.^12, 13^

### Physical Activity Time

Hours sleeping, sitting, and performing slight [e.g., standing, walking], moderate [e.g., household work], or heavy [e.g., heavy labor, intense exercise] activity during a typical 24-hour period were obtained through questionnaire administered during clinic visits, and were measured as a continuous score (Supplemental Table). Physical activity hours were weighted to reflect metabolic expenditure, calculated as: Physical Activity Score = 1.0*h_sleep_ + 1.1*h_sedentary_ + 1.5*h_slight_ + 2.4*h_moderate_ + 5.0*h_heavy_, with 24 hours representing the minimum possible score. For example, an individual who slept 8 hours, was sedentary for 9 hours, performed slight activity for 4 hours, moderate activity for 2 hours, and heavy activity for 1 hour had a weighted physical activity score of 33.7 hours for that day. This physical activity score has been validated and used previously.^14^

### Covariates

Participants also underwent a complete examination and completed additional questionnaires to obtain age, race/ethnicity, smoking history (current vs. not current and pack-years), weight (kilograms) and height (meters), which allowed us to calculate body mass index (BMI, kg/m^2^). Serum was also collected and used to measure RA-related autoantibodies, specifically rheumatoid factor (RF) measured by nephelometry, RF isotypes, IgM, IgG, and IgA, as well as anti-cyclic citrullinated peptide antibodies (anti-CCP2 and anti-CCP3.1) as described previously.^9^

### Statistical Analysis

We evaluated differences in demographics, autoantibody status, inflammatory markers, and physical activity time in FDRs with and without joint swelling or inflammatory joint symptoms using Student t-tests or Wilcoxon rank sum tests for continuous variables and chi-square tests or Fisher’s exact tests for categorical variables. We used a cross-sectional study design with analysis of covariance among 268 FDRs to examine whether baseline exam inflammatory joint symptoms were associated with baseline physical activity time. We then used a prospective study design to evaluate whether baseline joint swelling or symptoms were associated with physical activity over time, and whether changes in joint swelling or symptoms were associated with changes in physical activity over time. For the longitudinal analyses, we utilized linear mixed models, and adjusted for age, sex, race, BMI, smoking, and positivity for RA-related autoantibodies. To determine whether changes in joint symptoms were associated with changes in physical activity over time, we adjusted for baseline PA and incorporated an interaction term with time into the model, as it indicated whether there was an effect of time on the association between joint symptoms and physical activity hours, accounting for multiple visits per FDR. We also performed a sensitivity analysis for upper extremity joint swelling, stiffness, and pain, as well as for lower extremity joint swelling, stiffness, and pain following the above methods. All analyses were conducted in SAS version 9.4 (SAS Institute, Cary, North Carolina) and p-values < 0.05 were considered statistically significant.

## RESULTS

The 268 FDRs in this study had mean age of 50 ± 15 years at baseline, BMI of 28 ± 10 kg/m^2^, and a mean 37 ± 7 weighted physical activity hours per day. Table 1 presents participant characteristics, stratified by present joint swelling. At baseline, 20 (7%) FDRs had joint swelling, 50 (19%) reported joint stiffness, and 84 (31%) reported joint pain. These groups were not mutually exclusive with 11% reporting joint stiffness and pain, 3% with joint swelling and reporting pain, 0% with joint swelling and reporting stiffness, and 2% reporting all three – joint swelling, stiffness, and pain. FDRs with joint swelling, compared to FDRs without joint swelling, were older and were more likely to be anti-CCP2 and anti-CCP3.1 positive, and had slightly less physical activity time, a difference that did not achieve statistical significance. FDRs with joint stiffness and pain, however, had similar age, had similar autoantibody profiles, and had more physical activity time compared to FDRs without joint stiffness and pain (data not shown).

**Table 1.** Descriptive statistics by joint swelling at the baseline exam in 268 FDRs of patients with RA.

|  | **Joint Swelling**<br>**(n=20)** | **No Joint Swelling**<br>**(n=248)** | **p-value** |
| --- | --- | --- | --- |
| **Age, years (SD)** | 58 (17) | 49 (15) | 0.01 |
| **Female, %** | 60 | 71 | 0.28 |
| **Race, % Non-Hispanic White** | 90 | 81 | 0.54* |
| **BMI, kg/m<sup>2</sup>, mean (SD)** | 27 (6) | 28 (10) | 0.53 |
| **RF, % positive** | 10 | 14 | 1.00* |
| **CCP2, % positive** | 5 | 3 | 0.47* |
| **CCP3.1, % positive** | 17 | 9 | 0.33* |
| **RF IgM, % positive** | 0 | 13 | 0.08* |
| **RF IgG, % positive** | 10 | 28 | 0.11* |
| **RF IgA, % positive** | 10 | 3 | 0.17* |
| **Number of autoantibodies, mean (SD)** | 1 (1) | 1 (1) | 0.32 |
| **CRP (median, 25<sup>th</sup>, 75<sup>th</sup> percentile)** | 2 (1, 3) | 1 (1, 4) | 0.91** |
| **Pack-Years Smoking (median, 25<sup>th</sup>, 75<sup>th</sup> percentile)** | 0 (0, 8) | 0 (0, 4) | 0.26** |
| **Current Smoker, %** | 20 | 53 | 0.34* |
| **Physical Activity, hours, mean (SD)** |  |  |  |
| **Sleep** | 7 (1) | 7 (1) | 0.21 |
| **Sedentary** | 7 (3) | 7 (3) | 0.45 |
| **Light** | 5 (2) | 5 (3) | 0.25 |
| **Moderate** | 4 (3) | 3 (2) | 0.17 |
| **Heavy** | 1 (1) | 1 (2) | 0.17 |
| **Weighted Physical Activity Hours, mean (SD)** | 36 (5) | 37 (7) | 0.71 |

Values in this table are presented as means (and standard deviation) or as medians (and 25<sup>th</sup> and 75<sup>th</sup> percentile); and as proportions of FDRs with the specified characteristic. \*Indicates use of Fisher's exact test for p-values; and \*\*indicates use of Wilcoxon Rank Sum test for p-values.
RF = rheumatoid factor; CCP = cyclic citrullinated peptide, RF IgM = rheumatoid factor immunoglobulin M, RF IgG = rheumatoid factor immunoglobulin G, RF IgA = rheumatoid factor immunoglobulin

Cross-sectional analysis of inflammatory joint symptoms at baseline showed that after adjusting for age, sex, race, BMI, pack-years smoking, and positivity for RA-related autoantibodies, FDRs reporting joint pain had 1.3 more physical activity hours than FDRs without joint pain (38.1 vs 36.8 physical activity hours, respectively; p=0.15), although this association was not statistically significant. In contrast, FDRs self-reporting joint stiffness had 0.4 fewer physical activity hours than FDRs without joint stiffness (36.8 vs 37.3 physical activity hours, respectively; p=0.69); and FDRs with joint swelling documented by examination had 0.8 fewer physical activity hours than FDRs without joint swelling (36.4 vs 37.2 physical activity hours, respectively; p=0.60). Neither of these associations were statistically significant in fully adjusted analysis.

The average length of follow up was 1.2 years (range 14 days to 4.5 years). In this longitudinal analysis, adjusting for baseline physical activity time, age, sex, race, pack-years smoking, autoantibody status, and BMI, baseline joint symptoms were not significantly associated with a change in physical activity hours over time (Figure 1).

**Figure 1.**
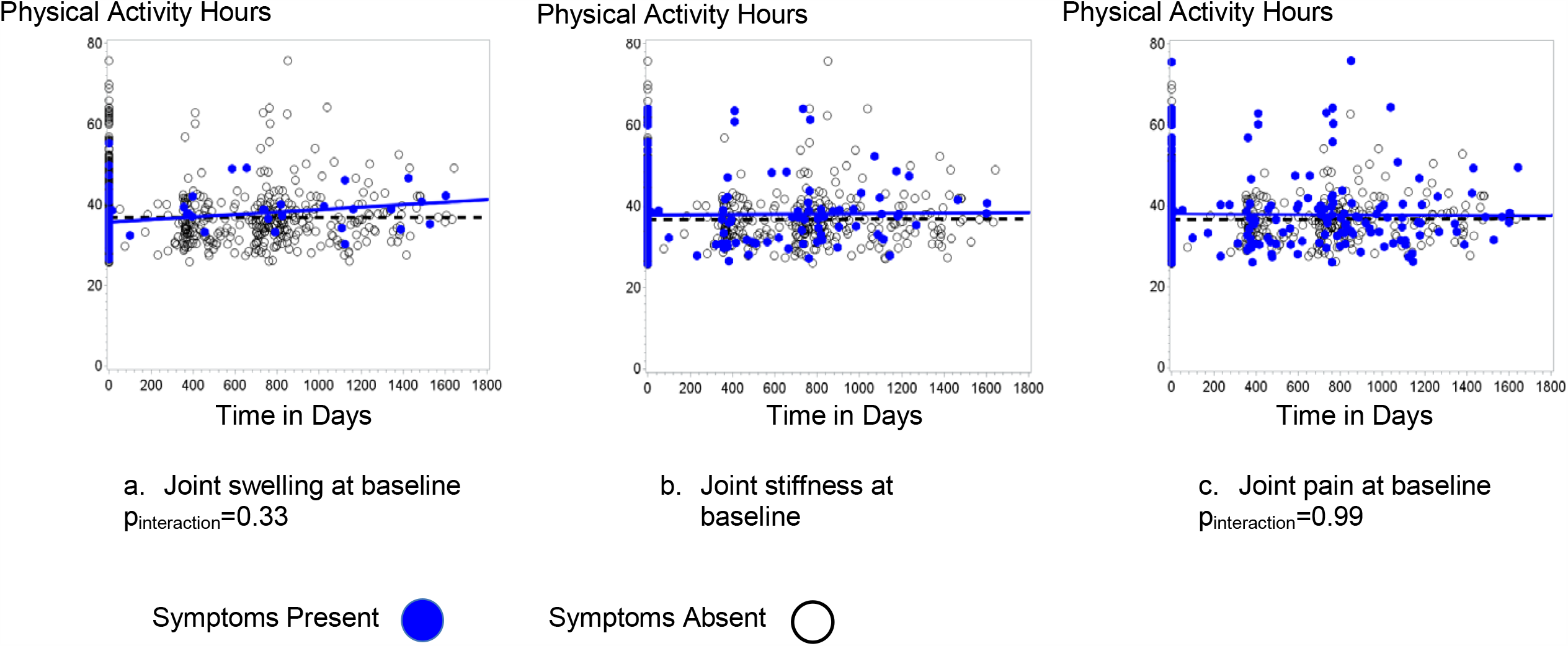
Associations between baseline joint swelling, stiffness, and pain, and physical activity over time in 268 FDRs of patients with RA. Figure 1 shows weighted physical activity hours over time in days for FDRs with joint swelling (1a), joint stiffness (1b), and joint pain (1c) at baseline, indicated by blue dots and blue solid lines, compared to FDRs without joint symptoms at baseline, indicated by black dots and black dashed lines. Analyses were fully adjusted for baseline physical activity hours, age, sex, race, pack-years of smoking, body mass index, and being positive for RA-related autoantibodies.

Longitudinal analysis evaluating the interaction between joint signs and symptoms and physical activity over time showed in fully adjusted models that FDRs with subsequent occurrences of joint swelling had more physical activity hours over time than FDRs without joint swelling (p_interaction_ <0.0001; β=0.004±0.001, p<0.0001). Conversely, FDRs reporting joint stiffness in subsequent clinic visits had fewer physical activity hours over time compared to FDRs who did not report joint stiffness (p_interaction_=0.0002; β=-0.001±0.001, p=0.11). Similarly, FDRs reporting joint pain in subsequent clinic visits had fewer physical activity hours over time compared to FDRs not reporting joint pain (p_interaction_=0.002; β=-0.001±0.001, p=0.32). (Figure 2)

**Figure 2.**
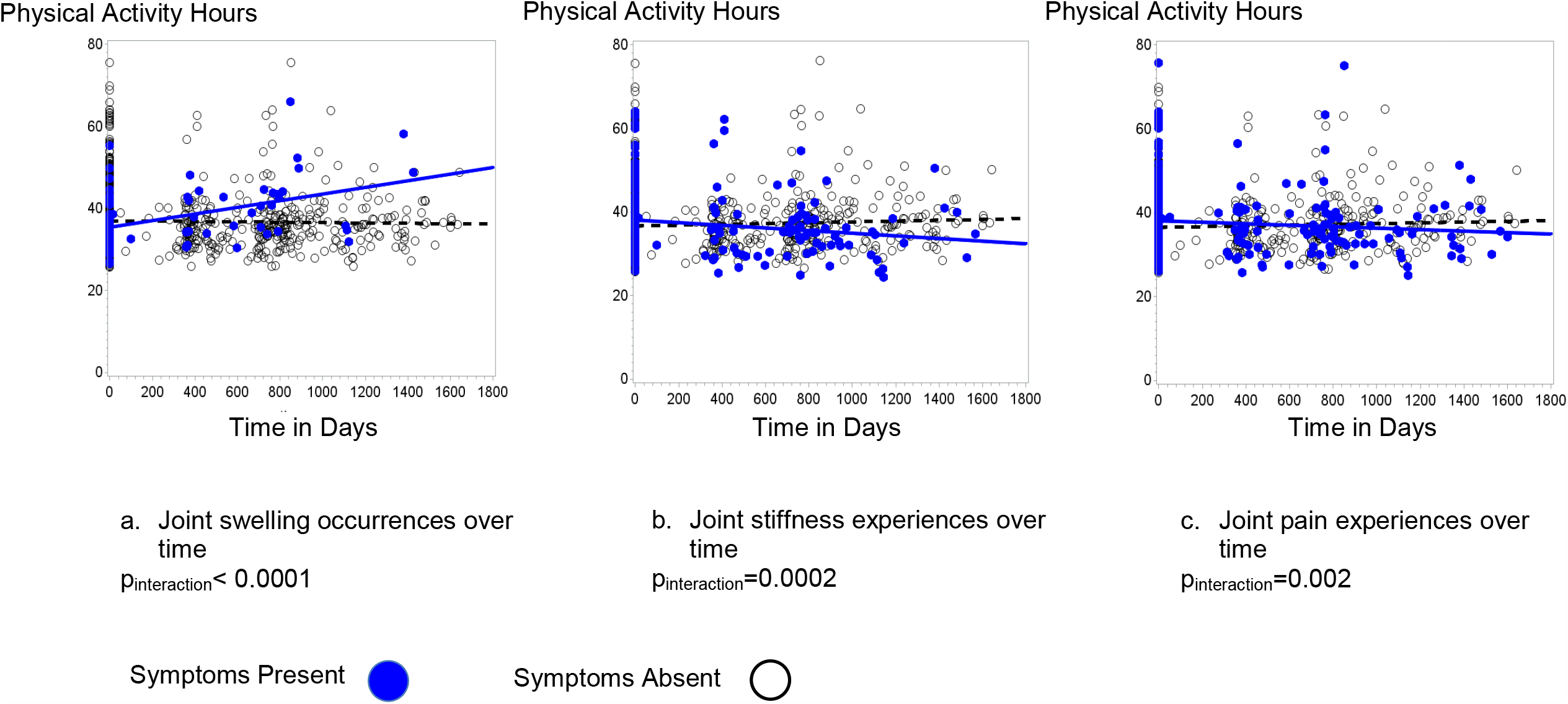
Associations between changes in joint swelling, stiffness, and pain, and physical activity over time in 268 FDRs of patients with RA. Figure 2 shows weighted physical activity hours over time in days for FDRs with joint swelling (2a), joint stiffness (2b), and joint pain (2c) during subsequent clinic visits, indicated by blue dots and blue solid lines, compared to FDRs without joint symptoms at subsequent clinic visits, indicated by black dots and black dashed lines. Analyses were fully adjusted for baseline physical activity hours, age, sex, race, pack-years of smoking, body mass index, and being positive for RA-related autoantibodies.

Findings from the sensitivity analysis for upper extremity joint swelling resembled the results from joint swelling, where in adjusted analysis, FDRs with upper extremity joint swelling had more physical activity hours over time than FDRs without upper extremity joint swelling (p_interaction_ <0.0001; β=0.003±0.001, p=0.003). Neither upper or lower extremity stiffness nor pain was associated with physical activity time in any of the sensitivity analyses (data not shown).

## DISCUSSION

In first-degree relatives of RA patients who were free of RA, we observed associations between joint swelling, joint stiffness, and joint pain and physical activity over time. Taking into account baseline physical activity hours, joint symptoms at baseline were not associated with physical activity hours over time. As FDRs developed joint symptoms over time, however, physical activity hours changed, where on average, FDRs reporting joint stiffness and pain had less physical activity, and FDRs experiencing joint swelling had more physical activity. These findings suggest current joint symptoms do not predict future physical activity, but that over time, average physical activity is influenced by joint symptoms. The inverse associations we found between joint stiffness and joint pain with physical activity are consistent with previous findings that RA flares and activity limitation are inversely associated with physical activity in RA patients;^2, 4, 5^ and indicate that similar joint involvement in a population without RA, but at higher risk for RA, may experience similar disease-specific barriers to physical activity as patients with active RA.

Physical activity is a robust predictor of multiple health outcomes such as diabetes, hypertension, and cardiovascular disease. Recent exciting and important findings suggest that physical activity prior to RA development has been associated with a lower risk of RA, a milder disease course, and less joint destruction.^6, 7^ In particular, increased time in self-reported physical activity 5 years before RA diagnosis was associated with decreased disease activity, joint pain, and inflammatory marker levels, as well as better self-reported health following disease onset.^7^ Despite these benefits, patients with RA have lower physical activity levels than their counterparts without RA.^2, 3^ It has been shown that RA patients average 1000 fewer steps per day during a flare period compared to a non-flare period.^4^ Yet among individuals who had established health-enhancing physical activity, their physical activity levels remained consistent even during periods of high RA disease activity.^15^ Given the modest results from our study, future directions for research in this population of FDRs include addressing joint stiffness and pain during this pre-clinical period. By addressing these symptoms, clinicians could potentially provide individuals at risk for RA the tools to establish and sustain regular physical activity prior to and potentially during the onset of RA; which could lead to the ability to maintain physical function and independent living during the course of the disease.

This study has important limitations. We were limited in our estimation of physical activity as particular tasks, e.g., walking versus rock climbing were not collected in this questionnaire. RA usually begins in the small joints of the hands and feet, and spreads later to the larger joints, as the articular cartilage and bone erode. Therefore, we chose to focus on the small joints of the hands and feet, as well as the elbow, in order to identify joint swelling and/or symptoms most indicative of earlier processes. In sensitivity analysis, however, we investigated whether upper extremity joint symptoms versus lower extremity joint symptoms were associated with physical activity hours. These findings were consistent with our primary findings, and suggest that physical activity hours did not differ based on whether joint symptoms were predominantly in the upper or lower extremities. The physical activity questionnaire captured activity on a ‘typical day’ and did not fully account for fluctuations of physical activity that could have been influenced temporarily by inflammatory joint signs and symptoms, illness, or other factors such as season of the year. Thus, our findings captured an overall picture of physical activity that likely underestimated associations between joint symptoms and physical activity time. Finally, because we investigated only FDRs of RA patients, we are not able to generalize these results to either patients living with RA, or to the general population.

This study is the first to our knowledge to investigate whether joint swelling or related symptoms are associated with physical activity in a population of FDRs of RA patients who do not have RA themselves, but have higher risk of RA. The associations between joint stiffness and pain and physical activity were consistent with our hypothesis and prior literature, whereas the association between joint swelling and physical activity was in a direction opposite to what we hypothesized. Future studies are needed to determine if symptoms of inflammatory arthritis are primary barriers to physical activity and whether intervening on the symptoms will allow continued pursuit of physical activity during this pre-clinical RA period.

In conclusion, among first-degree relatives of patients with RA, joint swelling, joint stiffness, and joint pain at baseline were not associated with future physical activity time. As joint swelling or symptoms changed over time, however, so did time in physical activity. Given these results, implications include adapting exercise and daily physical activity for individuals with joint pain or stiffness in order to intervene before the clinical manifestation of RA and potentially extend the period where they are RA-free.

## Data Availability

Data used for this analysis will be provided upon request.

## ACKNOWLEDGMENTS

The authors thank the SERA participants, investigators and staff for their valuable contributions to this research.

Funding for this research was made possible by grants from the Foundation for Physical Therapy Miami-Marquette Challenge Research Grant, the National Institute of Arthritis and Musculoskeletal and Skin Diseases (R01 AR051394-05), National Heart, Lung, and Blood Institute (K01HL122394), and National Institute for Diabetes and Digestive and Kidney Diseases (K24DK110427) at the National Institutes of Health, the ACR Rheumatology Research Foundation Within Our Reach: Finding a Cure for Rheumatoid Arthritis campaign and the Career Development Bridge Funding Award: K Bridge, and the Autoimmunity Prevention Center (U19 AI050864-10) at the National Institutes of Health.

## TABLES

**Supplemental Table.** Physical activity questionnaire with an example of reported values and resulting weighted physical activity score.

| Number of physical activity hours per day | Number of hours | Weight Factor | Example Weighted Hours |
| --- | --- | --- | --- |
| Sleep – Number of hours that you typically sleep? | 8 | 1.0 | 8 |
| Sedentary – Number of hours typically sitting? | 9 | 1.1 | 9.9 |
| Slight Activity – Number of hours with activities such as standing, walking? | 4 | 1.5 | 6 |
| Moderate Activity – Number of hours with activities such as housework | 2 | 2.4 | 4.8 |
| Heavy Activity – Number of hours with activities such as heavy household work, heavy yard work such as stacking or chopping wood, exercise such as intensive sports? | 1 | 5.0 | 5 |
| Total Number of hours (should be the total of above items) | 24 | Total Weighted Hours = 33.7 |  |
| Physical Activity Score = 1.0*h<sub>sleep</sub> + 1.1*h<sub>sedentary</sub> + 1.5*h<sub>slight</sub> + 2.4*h<sub>moderate</sub> + 5.0*h<sub>heavy</sub> |  |  |  |

## Notes

**FINANCIAL DISCLOSURES** The authors have no financial support from commercial sources or conflicts of interest to disclose.

### Competing Interest Statement

The authors have declared no competing interest.

### Author Declarations

University of California, San Diego Human Subjects Protection Program

